# Healthy behaviors and circadian patterns determined by actigraphy in researchers and administrative personnel as protective factors against metabolic disease and obesity

**DOI:** 10.1101/2022.12.07.22283230

**Authors:** Estefania Espitia-Bautista, Francisco Morales-Bautista, Ivette Rizo-Pastrana, Carolina Escobar, Christopher R. Stephens

## Abstract

**Objectives:** Three important, directly-causal, behavioral risk factors for obesity and its metabolic consequences are: food consumption, sedentary lifestyle and circadian disruption, such as social jet-lag, which should also partially explain the relevance of known, indirectly-causal, such as educational level, socio-economic status and/or type of job and its conditions. In this study we use actigraphy as a means to quantify and understand those differences in behaviors or conducts, related to 1) physical activity and 2) circadian disruption, which can explain observed differences in metabolic health in two different populations.

**Methods:** Metabolic and anthropometric data were taken from a population of university workers segmented by educational level – administrative workers (bachelor’s degree) and researchers (Masters or PhD degree) – Actigraphs collected temperature, acceleration, luxes and time in movement; participants use them for at least 1 week. Actigraphy data were divided in weekdays and weekends for analysis.

**Results:** We show that body mass index and metabolic syndrome criteria were significantly worse for the lower educational level group. Correspondingly, significant differences were found between administrative personnel and researchers across all measured actigraphy parameters – activity level, acceleration, light exposure and temperature. The most relevant differences are that researchers presented significantly more/less time in high/low-activity conducts, less differences in activity level between weekdays and weekends, and less social jet-lag than administrative personnel.

**Conclusions:** Researchers have healthier habits both in terms of voluntary physical activity and less circadian disruption, showing how work environment can be an important determinant of the degree to which healthy habits can be adopted.

**Highlights:**

- Educational level and work environment are relevant factors to contribute to the development of metabolic syndrome and obesity.
- Administrative personnel showed worse metabolic health than Researchers, and also Administrative workers showed less healthy behaviors in actigraphy.
- Differences between weekdays and weekends are a good parameter to measure social jet lag, but also for habits.
- Administrative personnel presented more Social jet lag than researchers, and more differences between weekdays and weekends in activity and temperature.

## INTRODUCTION

Obesity is one of the most prevalent diseases globally, although its etiology is complex and multi-factorial, it is, predominantly, a consequence of human behavior or conduct with two particularly important categories of conduct being those of food consumption and physical activity (PhyAct), which are associated with energy ingestion and expenditure [1,2].But it is been prove that circadian rhythms are relevant to the efficiency of an organism’s response to the daily challenges required by day–night cycles [3,4] and their long term disruption can cause a loss of homeostasis and lead to the development of diseases such as obesity [5,6].

An important factor that forms a nexus for obesity, habit formation and behaviour change is that of educational level (EduL). Many studies have shown that obesity [10,11], type-2 diabetes, hypertension and hyperlipidaemia [12] are more prevalent among those with lower educational attainment. Thus, one may posit that subjects with higher EduL have a greater ability to change habits, learning with adequate interventions and proving to have a higher degree of adherence to healthy behavior patterns [13,14].

Although objective measurement of those conducts that are most relevant for weight gain is challenging, actigraphy stands out as being particularly useful andcan be used to measure an important set of environmental and physiological variables, such as temperature [15], sleep duration [16], bedtime [17], exercise time [18], luxes and light-type during the day [19]and social jet-lag (SJL) [20], which is the discrepancy of the sleep midpoint between workdays and weekends [21]. All these factors have, in their turn, been related to the development of obesity and its consequences.

Given the important observed differences in weight and metabolic health as a function of EduL and/or type of job, in the present study we use actigraphy to infer and quantify some of the potentially different conducts that could be responsible for such differences. Specifically, we consider the results of an actigraphy study, as a function of EduL and, relatedly, of occupation, carried out in two population that have different habits because their schedule, with special emphasis on analyzing differences in circadian rhythms.

## 1. METHODS

### 2.1. Participants

#### 2.1.1. Study 1: Metabolic Syndrome

Data were taken from the “Project 42” database of Universidad Nacional Autonoma de Mexico (UNAM). We filtered two groups: Academics/Researchers (RM), who have a master’s degree or PhD (294 participants; 131 men and 163 women) and Administrative Personnel (APM), with high school diploma/bachelor’s degree (257 participants; 65 men and 192 women). We obtained self-reported hours of exercise per week, anthropometric measurements and biomarkers taken in fasting conditions to obtain metabolic syndrome (MS) criteria.

#### 2.1.2. Study 2: Actigraphy

Data were collected in 2019 in a new sample from UNAM. A cross-sectional study was performed using a group of 11 administrative personnel - APA – and a group of 15 researchers - RA. In this population, we used a bioimpedance scale to measure Body Mass Index (BMI), percentage of whole-body muscle and fat.

The project was approved by the Ethics Committee of the Facultad de Medicina, UNAM (FM/DI/152/2016). All participants were informed about the study’s purpose and data protection protocols:they signed a consent form.

### 2.2. Criteria for Metabolic Syndrome

MS has been defined by multiple associations [22–24], Here, we consider the following criteria: hyperglycemia (fasting blood glucose>100mg/dl); obesity, (BMI>30); waist circumference > 88cm in women and >102cm in men; High-Density Lipoprotein (HDL) <50 mg/dl in women and <40 mg/dl in men; hypertriglyceridemia (triglycerides >150 mg/dl); hypertension (>130 systolic and >85 diastolic), and insulin resistance in fasting (HOMA >3), calculated as (glucose mg/dl *insulin μU/ml)/405.

### 2.3. Ambulatory circadian monitoring (actigraphy)

Actigraphy was carried out consecutively for 6 weekdays and 1 or 2 weekends using the Kronowise® actigraph that monitor PhyAct, skin temperature and visible light intensity. All participants wear the device on their non-dominant wrist. The devices were configured to collect data every 30seconds. Data were separated into 24-hour segments by weekdays and weekend. SJL was calculated as Wittmann et al proposed in 2006. To test if there were similar changes to SJL using other variables, we used acrophases for time-above-threshold, temperature, light exposure (LEx) and acceleration to compare differences between weekdays and weekends. Additionally, we calculated parameters per day and per person and separated into weekdays and weekends (See parameters in detailed descriptions in Supplementary Table 1).

### 2.4. Statistics

Data are presented as mean ± standard error of the mean. MS data, SJL and all data obtained by Δ= (weekends - weekdays) were analyzed using Student-t tests. For the analysis of the circadian time-series we used a two-way Repeated Measures ANOVA for the factor “time” (hours) and the factor “groups” (APA/RA), or weekdays/weekends (“habits” or “routine”). Acrophases were obtained by a cosinor analysis in R [25]. Acrophases and parameters in actigraphy were evaluated with a two-way ANOVA for the factor groups or weekdays/weekends. We also used Pearson correlation and linear regression in acceleration vs temperature, high acceleration vs BMI and low acceleration vs BMI. Statistical analysis and graphs were elaborated using PRISM.6 (GraphPad Software).

## 3. RESULTS

### 3.1. Study 1: Anthropometry and metabolic biomarkers

In terms of MS, in Table 1 we see a comparison of averages, and in Supplementary Table2 (ST2) a comparison between the proportions of unhealthy and healthy levels in the two groups, as defined by the corresponding criteria for MS. For population averages, the statistically significant differences are in waist circumference, BMI and HDL (Table 1), with the RM group showing healthier levels. However, when we compare unhealthy/healthy proportions, as well as statistically significant differences in waist circumference, BMI and HDL, we also see significant differences in the HOMA index (ST2). We also note that the average per group results show that, for the APM group, their average exceeded the criteria for MS in both waist circumference (women) and triglycerides; while the RM group average was above the MS threshold only for triglycerides. Focus on the number of MS criteria, the average and proportions was significantly higher for the APM group (Table 1 and ST2). Moreover, from ST2 we note that 48.24% of the APM group satisfy at least 3 or more criteria for MS, while in the RM group only 27.89% reach 3 or more criteria, showing statistically significant differences between them.

**Table 1.** Metabolic syndrome criteria. Bold numbers indicates significant differences. Shaded cells indicates values that reached or surpassed metabolic syndrome criteria in each group.

| TABLE 1 | ADMINISTRATIVE PERSONNEL |  | RESEARCHERS |  |  |  |  |
| --- | --- | --- | --- | --- | --- | --- | --- |
|  | mean | sem | mean | sem | t | df | p |
| WAIST CIRCUMFERENCE MALE (>102 cm) | **99.24** | 1.37 | 95.59 | 0.77 | **2.49** | **194** | **0.0135** |
| WAIST CIRCUMFERENCE FEMALE (>88 cm) | **90.05** | 0.8 | 87.53 | 0.64 | **2.4** | **353** | **0.0168** |
| SYSTOLIC BP | 114.4 | 1.009 | 115.7 | 0.8929 | 0.96 | 549 | 0.33 |
| DIASTOLIC BP | 75.29 | 0.7149 | 76.31 | 0.6019 | 1.097 | 549 | 0.27 |
| IBM | **27.52** | **0.3327** | 26.07 | 0.2201 | **3.721** | **549** | **0.0002** |
| GLUCOSE | 98.05 | 1.915 | 96.16 | 1.7 | 0.741 | 549 | 0.45 |
| TRIGLYCERIDES | 166.6 | 6.307 | 161.8 | 5.665 | 0.567 | 549 | 0.57 |
| HDL MALE (<40 mg/dl) | **40.4** | **1.1** | 44 | 0.81 | **2.58** | **194** | **0.0106** |
| HDL FEMALE (<50 mg/dl) | **50.39** | **0.94** | 56.56 | 1.12 | **4.23** | **353** | **<0.0001** |
| HOMA | 2.302 | 0.1157 | 2.038 | 0.1027 | 1.717 | 549 | 0.086 |
| # CRITERIA MET SYN | **2.393** | **0.112** | 1.85 | 0.09315 | **3.75** | **549** | **0.0002** |
| H EXERCISE /WEEK | **2.218** | **0.2134** | 3.071 | 0.1662 | **3.194** | **549** | **0.001** |

We also found significant differences in weekly exercise hours, with the APM group showing less number of hours of weekly exercise (2.2) than the RM group (3.07) (See Table 1). Similarly, from the data shown in ST2, only 32.29% of the APM group exercise for more than 2 hours per week, while the corresponding figure for the RM group is 50.34%, with the differences being statistically significant (See ST2).

### 3.2. Study 2: Actigraphy

Actigraphy data were analyzed between groups, APA and RA, for weekdays and weekends respectively; and also for each group, contrasting weekdays versus weekends. Demographic data are shown in Supplementary Table 3. We note from Supplementary Figure 1 (SF1)A that the RA group has significantly more circadian movement towards the end of the day (20:00-0:00) on weekdays, whereas the APA group exhibited more activity at the beginning of the day (04:00-08:00). This pattern can be seen also in SF1G for the circadian acceleration measure, with an added difference that the average acceleration of the RA group was also greater between 16:00 and 20:00.

In terms of circadian temperature (SF1D), the RA group had a higher average temperature from 08:00-20:00. Furthermore, there was a significant difference in circadian LEx (SF1E) in the interval 08:00-12:00, with the RA group having significantly greater exposure. At weekends (SF1J, L, N, P), there were no significant differences for the interaction or the group factor except in the case of acceleration, where the RA group showed significantly higher values. (See Supplementary Table 4 (ST4) for more details).

Comparing weekdays and weekends for a given group, in SF1I, O we observe a significant decreased degree of activity for the APA group from 04:00-08:00 in both circadian movement and acceleration, while for the RA group the corresponding interaction effect was present only in the circadian movement factor. However, we observed a significant difference in the routine/habits factor for circadian acceleration of the RA group (SF1J, P). The APA group also showed a higher temperature from 08:00-16:00 and 04:00-08:00 on weekdays; while the RA group showed an increased temperature from 08:00-12:00, (SF1K, L). Although there were significant differences in the time factor for circadian LEx for the APA group, showing differences between day and night, there were no differences in the habits/routine factor or their interaction. In contrast, the RA group exhibited significant differences for interaction, for habits/routine and the time factors (SF1M, N; See ST4 for more statistical detail).

The acrophases on weekdays and weekends showed a difference ∼1 hour between the APA and RA groups on time-in-movement, showing significant differences for the groups factor, but not the routine/habits factor or their interaction (Figure 1A). Acceleration showed a similar pattern, with a difference ∼1 hour between the APA and RA groups, and a significant difference only for the groups factor (Figure 1D). No significant differences were observed at weekends for any factor. For temperature or LEx acrophases, we found no significant differences for any factor (Figure 1B, C; See ST4 for more statistical details).

**Fig. 1.**
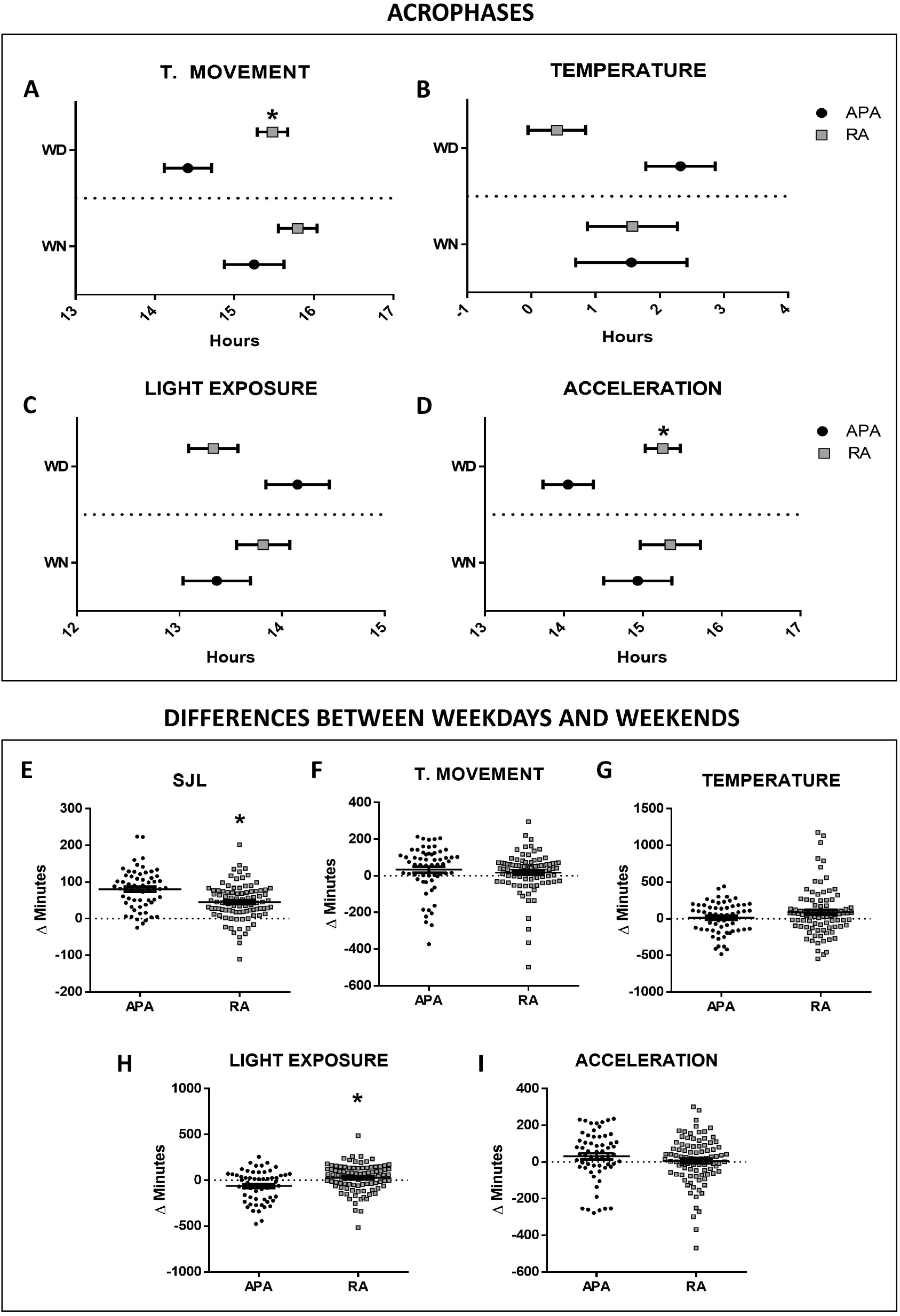
From A to D: Acrophases on Weekdays (WD) and Weekends (WN) in Administrative Personnel (APA) and Researchers (RA). (A) Total Time Above Threshold (T. Movement). (B) Temperature. (C) Light exposure. (D) Acceleration. Data are shown as mean ± s.e.m. (*) indicates a significant difference between the groups (p < 0.05). From E to I: Differences in Administrative Personnel (APA) and Researchers (RA) between weekdays and weekends (Δ=WN-WD), obtained from the acrophases for each variable. (E) Social Jet Lag. (F) Total Time Above Threshold (T. Movement). (G) Temperature. (H) Light exposure. (I) Acceleration. Data are shown as mean ± s.e.m. (*) indicates a significant difference between the groups (p < 0.05)

For SJL, we see that the RA group have significantly less minutes of difference between weekdays and weekends than the APA group (Figure 1E). Using acrophases, we also calculated the differences between weekdays and weekends, observing significant differences only in LEx, where APA showed a difference ∼ - 60 min, while for the RA group the difference was only ∼25min (Figure 1H). Movement, acceleration and temperature showed no significant differences (Figure 1F, G, I; See ST4 for more statistical details).

When we evaluate the total activity per day on weekdays and weekends, we observed significant differences in the routine/habits factor (Table 2 (T2) A and ST4). We also note that at weekends the RA group exhibits higher levels of acceleration than the APA group, with significant differences in the group factor for both average acceleration and AUC (T2B, C and ST4). We divided the day into activity and rest periods for each person based on the acceleration variable, observing significant differences only at weekends, with the RA group showing higher activity than the APA (T2D, E and ST4).We also considered further differences by dividing the activity period into high activity and low activity, observing that the RA group spent more time in high activity, on both weekdays and weekends, than the APA group (T2F, G and ST4).

**Table 2.** Activity parameters presented as mean +/- sem on weekdays and weekends. (*) indicates significant difference from Administrative Personnel and (^#^) indicates significant difference from Weekdays (p<0.05).

| Table 2 |  | WEEKDAYS |  |  |  | WEEKENDS |  |  |  |
| --- | --- | --- | --- | --- | --- | --- | --- | --- | --- |
|  |  | ADMINISTRATIVE PERSONNEL |  | RESEARCHERS |  | ADMINISTRATIVE PERSONNEL |  | RESEARCHERS |  |
|  |  | mean | sem | mean | sem | mean | sem | mean | sem |
| A | TOTAL ACTIVITY (24H-T.MOV) | 296343.8 | 7202.15 | 290897.4 | 3970.05 | 269134.5 | 11455.41 | 276148.9 | 9486.64 |
| B | TOTAL ACTIVITY (24H-A-G) | 12.34 | 2.48 | 13.38 | 3.56 | 10.8 | 1.95 | **13.16\*** | **3.76** |
| C | AUC (A-G) | 17743.28 | 442.68 | 18908.27 | 421.06 | 15551.07 | 627.78 | **18772.73\*** | **921.99** |
| D | ACTIVITY PERIOD (A-G) | 16.71 | 3.29 | 17.82 | 4.41 | 15.65 | 2.19 | **18.72\*** | **5.83** |
| E | REST PERIOD (A-G) | 2.43 | 0.07 | 2.56 | 0.05 | 2.22 | 0.12 | 2.57 | 0.09 |
| F | HIGH ACTIVITY (MIN A-G) >80 | 6.81 | 1.49 | **21.24\*** | **2.7** | 4.4 | 2.06 | **21.63\*** | **5.1** |
| G | LOW ACTIVITY (MIN A-G) <5 | 244.15 | 11.16 | 221.14 | 8.23 | 250.27 | 16.28 | 212.66 | 13.77 |
| H | BED TIME | 23:06 | 0.15 | **0:02\*** | **0.14** | **0:03<sup>#</sup>** | **0.28** | 0:28 | 0.25 |
| I | WAKE UP TIME | 5:54 | 0.15 | **7:00\*** | **0.12** | **7:42<sup>#</sup>** | **0.3** | **8:07<sup>#</sup>** | **0.24** |
| J | RESTING (MIN) | 444.4 | 9.94 | 431.2 | 8.13 | **526.4<sup>#</sup>** | **24.79** | **493.01<sup>#</sup>** | **22.48** |
| K | LUXES IN ACTIVITY PERIOD | 346.07 | 39.34 | 407.76 | 29.97 | 455.4 | 73.33 | **688.71<sup>#</sup>** | **127.75** |
| L | LIGHT AT NIGHT (MIN) >5 (12H) | 171.17 | 9.17 | **208.24\*** | **9.6** | 122.92 | 15.46 | 172.69 | 15.42 |
| M | TEMPERATURE 24H | 32.36 | 0.07 | **32.77\*** | **0.07** | 32.31 | 0.15 | 32.62 | 0.15 |
| N | TEMPERATURE AT DAY | 31.72 | 0.1 | **32.38\*** | **0.08** | **32.30<sup>#</sup>** | **0.14** | 32.62 | 0.15 |
| O | TEMPERATURE AT NIGHT | 33.8 | 0.07 | 33.64 | 0.08 | 33.89 | 0.16 | 33.81 | 0.13 |

We also found that the RA group had a bedtime ∼1 hour later than the APA group on weekdays, but with no difference at the weekends (T2Hand ST4). Similarly, the RA group wake up ∼1 hour later than the APA group on weekdays; while at weekends both groups wake up later than on weekdays (T2I and ST4). In addition, we estimated the total resting time and found that both groups rest more at the weekends (T2J and ST4).

LEx was also evaluated in the activity period, with the result that the RA group showed more luxes at weekends than on weekdays (T2K and ST4). For minutes exposed to light at night (>5 lux) we found that the RA group spends more time at night with luxes >5 on weekdays, finding differences in the group factor and the routine/habits factor (T2L and ST4).

Finally, higher 24-hour averaged temperature levels were observed for the RA group on weekdays, but with no differences at weekends (T2M and ST4). The same difference in averages was also noted during the day (activity), but now with the APA group exhibiting higher temperatures at the weekends. At night no significant differences were observed (T2N and O; and ST4).

As a further analysis, we considered the correlation between temperature and acceleration in the activity period (09:00 to 21:00) as activity s reveal periods of intense activity, such as exercise, with a consequent temperature regulation. We observed that for both groups, both on weekdays and weekends, the relation between acceleration and temperature is negative (Figure 2 A-D). However, as seen in Figure 2, the RA group’s distribution exhibited a much more extensive “tail” towards higher accelerations (from ∼ 100-320) on both weekdays and weekends.

**Fig. 2.**
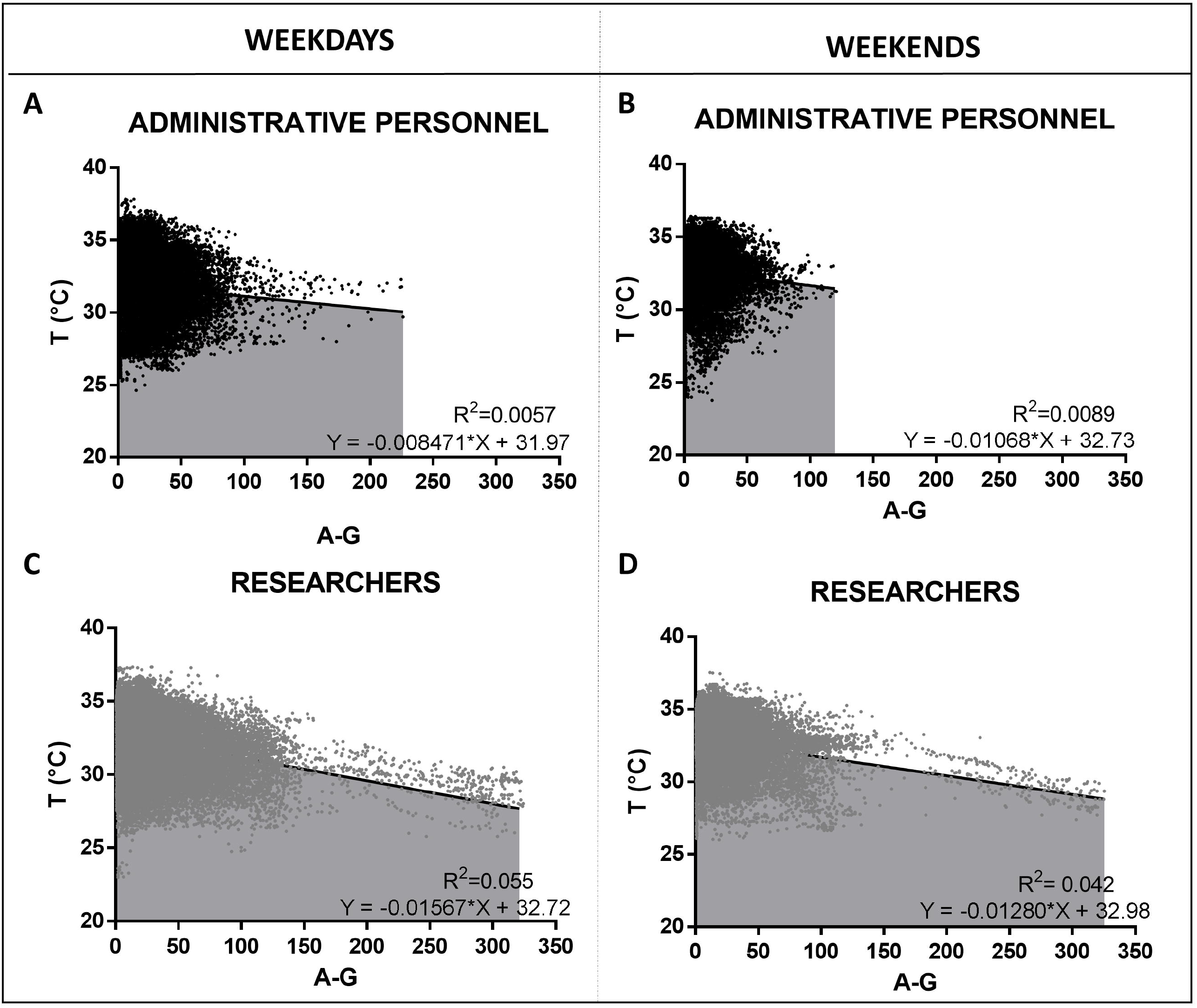
Correlation between Acceleration (A-G) and Temperature (T°C) using each individual measurement on weekdays (left; A and C) and at weekends (right; B and D) obtained from Administrative Personnel and Researchers. Black lines are linear regressions of the relation with their associated equation and R^2^. The Pearson correlation coefficient for each analysis, with r= -0.076 for the APA group on weekdays, while at weekends r= -0.095; in contrast, r= -0.236 for the RA group on weekdays and r= -0.205 at weekends.

Finally, when we correlate each day of weekdays or weekends of low activity vs BMI, we observe in Figure 3 (A and B) that the APA group exhibit higher positive correlations than the RA group (C and D). The Pearson correlations on weekdays for APA were r=0.34 (p=0.0043), and r=0.39 (p=0.08) at weekend, while the RA group on weekdays had r=-0.13 (p=0.19), and r= 0.16 (p=0.32) at weekends. In contrast, for the RA group there was no significant deviation from zero for either regression. On the contrary, when we correlate high activity vs BMI, we observe (Figure 3E and F) that the RA group now exhibits a significant negative relation between BMI and minutes spent in high activity, with a regression coefficient of -2.3 (p=0.0005, r=-0.13) on weekdays. On the contrary, for the APA group there is no significant linear relation.

**Fig. 3.**
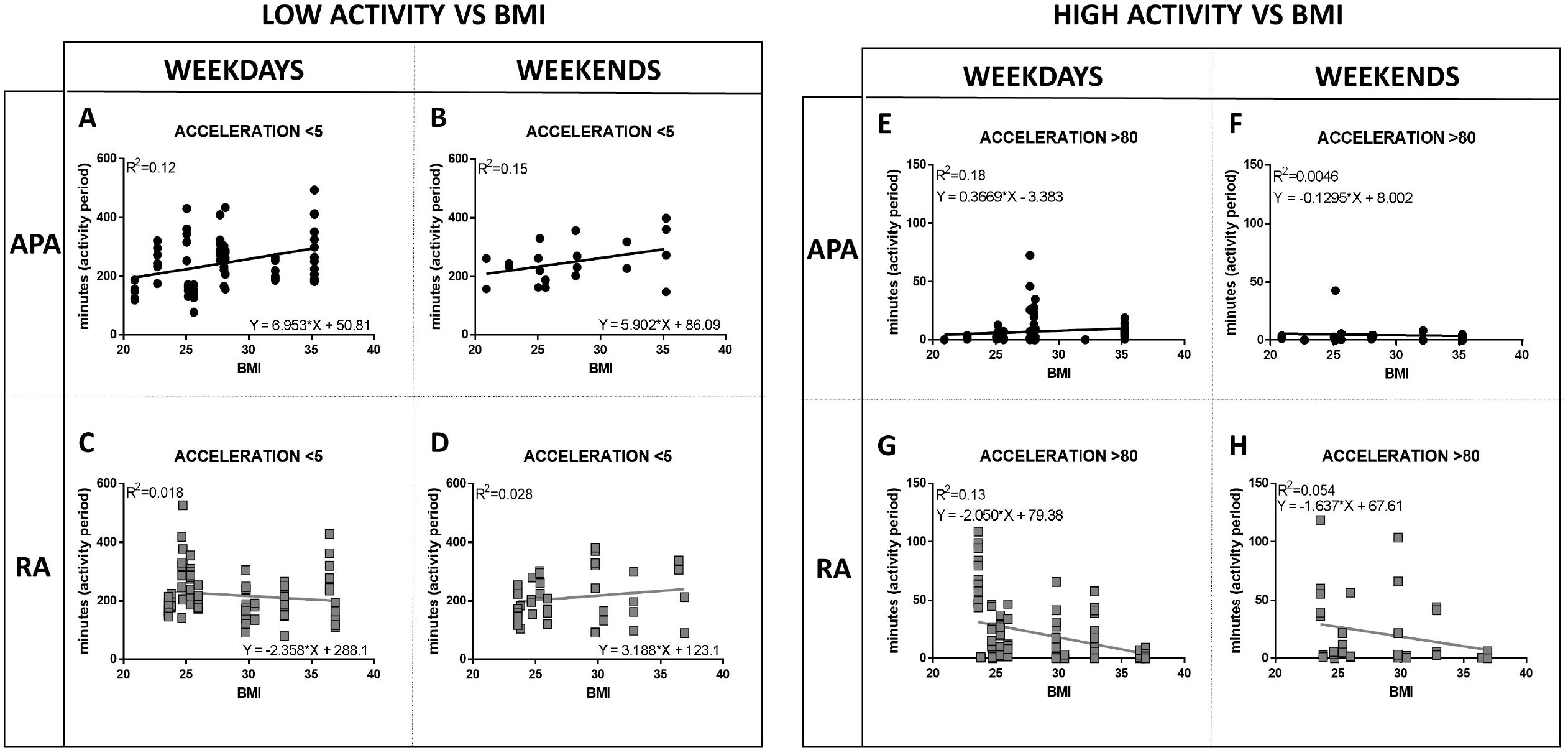
From A to D: Correlation between low acceleration (<5) and Body Mass Index (BMI) using every day from each participant on weekdays (left; A and C) and on weekends (right; B and D), obtained from Administrative Personnel (A and B, in black circles) and Researchers (C and D, in grey squares). Data were taken only from activity period. Black lines are linear regressions of the relation with their associated equation and R^2.^ From E to H: Correlation between high acceleration (<80) and Body Mass Index (BMI) using every day from each participant on weekdays (left; E and G) and on weekends (right; F and H), obtained from Administrative Personnel (E and F, in black circles) and Researchers (G and H, in grey squares). Data were taken only from activity period. Black lines are linear regressions of the relation with their associated equation and R^2^

## 4. DISCUSSION

Although the factors that cause obesity and metabolic disease range from the “micro” to the “macro”, it is conduct that forms the nexus between physiology and environment. As it is well established that higher EduL generally correlates with better metabolic health [11,26], we should ask how EduL is reflected in energy balance? Also, PhyAct is an important component of our circadian rhythms, and their disruption, has also been identified as an important risk factor for obesity [6]. Here, we used actigraphy to investigate the temporal distribution and the intensity of PhyAct, as well as identify and analyze differences in circadian rhythms, as factors that may explain differences in health outcomes as a function of EduL related with the schedule of each population.

There are important differences in metabolic health as a function of EduL in our study population, with the RM group having significantly lower average BMI and higher levels of HDL. Most striking is the number of MS criteria associated with each group. In terms of lifestyle factors, we noted that the RM group reported a significantly greater number of hours of exercise per week. Although the relation between EduL and metabolic health has been amply discussed, higher education levels – Bachelors, Masters and Doctorate – have been grouped together [26,27]. Here, as was shown previously [11] we see that there are also significant health differences between graduate level participants have worse metabolic health than their postgraduate counterparts, and here it is important to notice that their jobs are completely different.

As self-reported exercise is subject to reporting bias, a goal of the actigraphy study was to determine if there were any measurable objective behavioral or environmental factors that could partially explain the significant health differences between these two groups. Our results clearly indicate two distinct categories of such differences: PhyAct and disruption of circadian rhythms. For activity, on weekdays, the APA group are physically active before the RA group who, in turn, exhibit much more activity, and especially, relatively intense activity, towards the end of the workday. This may reflect the more liberal work regime that researchers enjoy relative to their administrative counterparts, a difference that is also reflected in both their bedtime and wakeup times, with the APA going to bed an hour earlier than the RA and waking up an hour earlier. However, at the weekends when there is no schedule for work, both groups go to bed and wakeup somewhat later; but, the SJL associated with the APA group is more than an hour (∼80 min) and is significantly higher than the RA group who have a SJL of only 44 min. Previous results have shown that people with more than 2 hours of SJL have a higher risk of developing metabolic and weight problems, as well as presenting higher cortisol, less PhyAct and a higher resting heart rate [28,29].

Although during weekdays there is an important difference between APA and RA in terms of the temporal distribution of their activity, at the weekends (SF1B), this difference is absent. However, there is a large difference in the intensity of the activity, as seen in SF1H. This difference is also present during the week in the late afternoon and evening. Further evidence for the more intense activity patterns of the RA group can be gleaned from T2G, where we see that the RA group spend more time in high intensity activity (acceleration>80) both on weekdays and at weekends, with the average difference ∼15 min. Although, at first sight, this may seem an insignificant difference, it represents a 300-500% increase for the RA group over the APA group.

An important characteristic of obesity is that it is a result of small changes over long time periods rather than large changes over short periods [30,31]. It’s been reported that moderate exercise for 15 minutes could lead to a calorie burn of ∼ 50-100 calories, which, over long periods, could lead to a substantial difference [32]. The fact that the RA group is associated with higher levels of intensity of PhyAct is also consistent with the relation of self-reported hours of exercise between the RM and APM groups. Also, the APA group had significantly longer low-activity periods (acceleration<5), reinforcing the conclusion that the RA group has higher levels of PhyAct across a spectrum of activity levels. As seen in Figure 3, these differences in time spent in low versus high activity levels are related to the BMI distribution of the APA and RA groups, with the BMI of the latter being significantly lower/higher for those Researchers with higher/lower high-activity level. Similarly, the BMI of the APA group is higher/lower for those Administrative Personnel with significantly higher/lower low-activity levels. We hypothesize that, just as a relatively small number of minutes spent in high activity can have a significant beneficial effect on BMI, so a relatively small number of “excess” minutes spent in low activity can have a detrimental effect.

We interpret the overall activity patterns of PhyAct, as reflecting stronger habit formation in Researchers with respect to high PhyAct that is reflected in both their work-day routine and at weekends, resulting in better metabolic health. Additionally, although both groups change certain habits at the weekends – bedtime, wake time and rest time – it is the APA group that exhibits the bigger changes, as is manifest in their greater SJL, which we take to be associated with a desire to be less constricted with respect to their routines, particularly with their wakeup times. Although during weekdays one could argue that the more restrictive work schedule of the Administrative Personnel makes it more difficult to exercise, often they work till 19:00-20:00, there is no such restriction at the weekends. Rather, it is the continuation of a *relatively* unhealthy exercise habit that is continued into the weekend. Studies have shown that differences in SJL are important for metabolic health [28]. However, these studies only refer to the discrepancy in the sleep time between weekdays and weekends. We believe it is important to emphasize that the changes between weekdays and weekends involve more than just sleep. We have demonstrated that besides changes in sleep time, there are significant changes in the type and quantity of activity (high or low), LEx, resting and temperature; all of which are potentially important factors that reflect conducts related to the development of obesity.

With respect to LEx, we observed that RA are exposed to higher levels than APA, and at weekends showed significantly higher levels than on weekdays, which may indicate more time spent outside at weekends, while the APA group spend more time indoors resting. Several studies have shown that LEx, in the correct phase, helps to maintain circadian rhythms and is also associated with better mood states [33,34]. Analyzing light at night, which is related to the development of overweight and metabolic problems [35,36], we only observed differences on weekdays, with RA showing more minutes with light at night. This can be explained by the later bed time for the RA group.

With respect to temperature, it is noteworthy that the RA group is associated with significantly higher temperatures than the APA group from 08:00-20:00 on weekdays. Of course, one could try to interpret this in terms of physiological differences between the two groups, as in Figure 2, where the higher degree of intense PhyAct of the RA group is associated with lower temperatures, potentially due to a cooling response. However, this would lead to a lower average temperature for this group, not a higher one. Several studies have shown that people with obesity show higher mean temperature than lean people [37] reflecting how people with obesity offset excess calorie intake by a higher heat transfer to the environment. However, others have claimed that people with obesity present lower temperatures and therefore accumulate more fat because they dissipate less excess calories as heat [38].

There are, of course, several limitations in our work. Firstly, the metabolic data and the actigraphy data were taken from two distinct samples. We believe, however, that both datasets are representative of the overall population. A second point is that there may exist confounding factors for the differences we have observed other than EduL, the type of job and their schedule, or even the environmental work conditions. An important one, which is correlated to EduL in this case, is the occupation of the participants–, their associated socio-economic status and age. Certainly, occupation is related to both work environment and daily routine. As mentioned, Administrative Personnel have less freedom to come and go as compared to Researchers, as well as more fixed working hours.

## 5. CONCLUSIONS

EduL has been reported as a relevant factor in the incidence of obesity and metabolic disease. However, they are not themselves *directly* causal. In this paper, having previously determined the importance of EduL, beyond the graduate level, as a predictor of obesity and metabolic disease, we use actigraphy data to determine if there were differences in conduct between Administrative Personnel and Researchers that could explain their quite distinct health outcomes. Although actigraphy data is subtle to interpret, it can also be very revealing. We determined that consistent with their healthier metabolic state and greater self-reported exercise hours, Researchers were associated with healthier habits associated with both higher intense PhyAct levels, both during weekdays and at the weekends, as well as with less SJL.

## Data Availability

All data produced in the present study are available upon reasonable request to the authors

https://chilam.c3.unam.mx/proy-conductome/datos-conductome

## 6. ACKNOWLEDGEMENTS

We thank Romel Calero and the rest of the Project 42 team for data collection.

## 9. DATA AVAILABILITY STATEMENT

The datasets generated during and/or analysed during the current study are available in https://chilam.c3.unam.mx/proy-conductome/datos-conductome

## 10. AUTHORS CONTRIBUTION

**EEB:** Conceptualization, design of the study, supervision, analysis and interpretation data, manuscript writing.

**FMB:** Analysis data and manuscript writing

**IRP:** Conduct of the study, data collection and manuscript writing.

**CE:** Analysis and interpretation data, manuscript writing (review and editing)

**CRS:** Conceptualization, funding acquisition, supervision, design of the study, interpretation data, manuscript writing (review and editing)

## 14. SUPPLEMENTARY FIGURES

**Supplementary Figure 1**: From A to H: Circadian differences between Researchers (RA, grey lines) and Administrative Personnel (APA, black lines) on weekdays (left) and weekends (right). (A and B) Circadian movement as a function of time: total Time Above Threshold (T.MOV). (C and D) Circadian temperature in centigrade (C°) as a function of time. (E and F) Circadian light exposure as a function of time in luxes. (G and H) Circadian acceleration as a function of time in standard acceleration (A-G). Data are shown as mean ± s.e.m. (*) indicates significant difference between the groups at each temporal point (p < 0.05). From I to P: Circadian differences between Weekdays (WD, straight line) and Weekends (WN, dotted line) in Administrative Personnel (left, black lines) and Researchers (right, grey lines). (I and J) Circadian movement as a function of time: total Time Above Threshold (T.MOV). (K and L) Circadian temperature in centigrade (C°) as a function of time. (M and N) Circadian light exposure as a function of time in luxes. (O and P) Circadian acceleration as a function of time in standard acceleration (A-G). Data are shown as mean ± s.e.m. (*) indicates significant difference between weekday and weekend at each temporal point (p < 0.05)

