## supplementary table 1 for "Healthy behaviors and circadian patterns determined by actigraphy in researchers and administrative personnel as protective factors against metabolic disease and obesity"

Supplementary Table 1. Description of each parameter used in actigraphy per day, per person.

| **PARAMETER** | **DESCRIPTION** |
| --- | --- |
| **Total activity (24hours)** | Sum of all counts across 24 hours of recording (time above threshold) |
| **Total activity (24hours)** | Average acceleration across a 24-hour period. |
| **Area under the curve (AUC)** | Using data from acceleration in 24-hour. |
| **Activity period (acceleration during the day)** | Average in acceleration at day, beginning when counts start upon 100 in 5 consecutive count, in time above threshold, after rest period (counts in 0 at night), without temperature dropping, until rest period. |
| **Rest period (acceleration at night)** | Average in acceleration at night, beginning when counts in 0 were presented for 10 consecutive periods in time above threshold and maintained until activity period started. |
| **Minutes spend in high activity** | Number of counts in acceleration >80, divide in 2 to obtain the minutes. |
| **Minutes spend in low activity** | In order to avoid register low activity because people are asleep at night, we only calculate in the activity period the number of counts in acceleration between 0 and 5, divide in 2 to obtain the minutes. |
| **Bedtime** | Identified when the rest period started. To avoid the change at midnight, we considered midnight=0, in this way 23:00 = -1, while 1:00 = 1. |
| **Wake up time** | Identified when activity period started. |
| **Minutes resting** | Time in minutes identified between bedtime and wake up time, adding the minutes of inactivity periods during the day. |
| **Inactivity periods during the day** | Identified when 0 was presented more than 5 consecutive times in activity period, but the temperature was maintained and did not dropped, meaning that the person was not asleep). |
| **Luxes in the activity period** | Average taken from light exposure during activity period. |
| **Light at night (12-hours)** | Minutes from 21:00h to 9:00h with luxes >5. |
| **Temperature 24-hours** | Average taken across 24 hour. |
| **Temperature during the day** | Average taken across activity period. |
| **Temperature at night** | Average taken across rest period. |
