## supplementary table 2 for "Healthy behaviors and circadian patterns determined by actigraphy in researchers and administrative personnel as protective factors against metabolic disease and obesity"

Supplementary Table 2. Proportion of the population associated with a particular Metabolic Syndrome criterion. Bold numbers indicates significant differences.

| **Supplementary Table 2** | **ADMINISTRATIVE PERSONNEL** | | **RESEARCHERS** | | Chi-square test | Fisher´s exact test |
| --- | --- | --- | --- | --- | --- | --- |
|  | % reached criteria | % healthy | % reached criteria | % healthy |  |  |
| **WAIST CIRCUMFERENCE** | **55.25** | **44.75** | 43.19 | 56.81 | **7.97** | **0.0049** |
| **BLOOD PRESSURE** | 17.89 | 82.11 | 15.3 | 84.7 | 3.4 | 0.082 |
| **BMI** | **24.9** | **75.1** | 15.98 | 84.02 | **6.77** | **0.0106** |
| **GLUCOSE** | 26.45 | 73.55 | 21.08 | 78.92 | 2.19 | 0.159 |
| **TRIGLYCERIDES** | 44.74 | 55.26 | 44.21 | 55.79 | 0.015 | 0.931 |
| **HDL** | **51.75** | **48.25** | 35.03 | 64.97 | **15.65** | **<0.0001** |
| **HOMA** | **22.95** | **77.05** | 15.98 | 84.02 | **4.28** | **0.042** |
| **# CRITERIA MET SYN** | **48.24** | **51.76** | 27.89 | 72.11 | **24.28** | **<0.0001** |
| **H EXERCISE /WEEK** | **67.71** | **32.29** | 49.66 | 50.34 | **18.34** | **<0.0001** |
