## supplementary table 3 for "Healthy behaviors and circadian patterns determined by actigraphy in researchers and administrative personnel as protective factors against metabolic disease and obesity"

Supplementary Table 3. Demographic data for Actigraphy presented as mean +/- sem. Bold numbers indicates significant differences.

|  | **ACTIGRAPHY** | | | |  |  |  |
| --- | --- | --- | --- | --- | --- | --- | --- |
| **Supplementary Table 3** | **ADMINISTRATIVE PERSONNEL (n=11)** | | **RESEARCHERS (n=15)** | | t | df | p |
|  | mean | sem | mean | sem |  |  |  |
| **AGE (years)** | **52.73** | **1.75** | 45.33 | 2.35 | **2.35** | **24** | **0.02** |
| **HEIGHT (cm)** | 156.5 | 2.53 | **168.7** | **2.51** | **3.33** | **24** | **0.002** |
| **WEIGHT (kg)** | 67.78 | 3.12 | **80.55** | **3.46** | **2.62** | **24** | **0.014** |
| **BMI** | 27.81 | 1.42 | 28.41 | 1.2 | 0.32 | 24 | 0.74 |
| **MUSCLE%** | 24.1 | 0.81 | **30.61** | **1.49** | **3.44** | **24** | **0.0021** |
| **FAT%** | **42.51** | **2.12** | 32.39 | 2.74 | **2.74** | **24** | **0.011** |
