## supplementary table 4 for "Healthy behaviors and circadian patterns determined by actigraphy in researchers and administrative personnel as protective factors against metabolic disease and obesity"

Supplementary Table 4. Statistical data from Figure 1, Supplementary Figure1; and Table 2. Bold numbers indicates significant differences.

| **Supplementary Table 4** | **TWO-WAY ANOVA** | | | | | | | | | |
| --- | --- | --- | --- | --- | --- | --- | --- | --- | --- | --- |
| **Supplementary Figure 1** | **INTERACTION** | | | | **GROUP FACTOR** | | | **TIME ALONG THE DAY** | | |
|  | F | | p | | F | p | | F | p | |
| **(A) CIRC. T. MOV. WD** | **F (5, 770) = 16.75** | | **P < 0.0001** | | F (1, 154) = 0.46 | P = 0.49 | | **F (5, 770) = 404.9** | **P < 0.0001** | |
| **(B) CIRC. T. MOV. WN** | F (5, 270) = 0.90 | | P = 0.47 | | F (1, 54) = 0.32 | P = 0.56 | | **F (5, 270) = 119.5** | **P < 0.0001** | |
| **(C) CIRC. LIGHT EXP. WD** | **F (5, 770) = 5.93** | | **P < 0.0001** | | F (1, 154) = 2.08 | P = 0.15 | | **F (5, 770) = 63.54** | **P < 0.0001** | |
| **(D) CIRC. LIGHT EXP. WN** | F (5, 270) = 0.72 | | P = 0.60 | | F (1, 54) = 1.86 | P = 0.17 | | **F (5, 270) = 14.77** | **P < 0.0001** | |
| **(E) CIRC. TEMP. WD** | **F (5, 770) = 7.43** | | **P < 0.0001** | | **F (1, 154) = 14.34** | **P = 0.0002** | | **F (5, 770) = 125.5** | **P < 0.0001** | |
| **(F) CIRC. TEMP. WN** | F (5, 270) = 0.62 | | P = 0.67 | | F (1, 54) = 0.38 | P = 0.53 | | **F (5, 270) = 26.06** | **P < 0.0001** | |
| **(G) CIRC. ACCELERATION WD** | **F (5, 770) = 13.87** | | **P < 0.0001** | | F (1, 154) = 2.99 | P = 0.08 | | **F (5, 770) = 176.4** | **P < 0.0001** | |
| **(H) CIRC. ACCELERATION WN** | F (5, 270) = 0.57 | | P = 0.72 | | **F (1, 54) = 7.24** | **P = 0.009** | | **F (5, 270) = 36.56** | **P < 0.0001** | |
| **Supplementary Figure 1** | **INTERACTION** | | | | **ROUTIN/HABITS FACTOR** | | | **TIME ALONG THE DAY** | | |
|  | F | | p | | F | p | | F | p | |
| **(I) CIRC. T. MOV. APA** | **F (5, 420) = 10.57** | | **P < 0.0001** | | F (1, 84) = 3.50 | P = 0.06 | | **F (5, 420) = 143.1** | **P < 0.0001** | |
| **(J) CIRC. T. MOV. RA** | **F (5, 620) = 4.62** | | **P = 0.0004** | | F (1, 124) = 2.24 | P = 0.13 | | **F (5, 620) = 289.3** | **P < 0.0001** | |
| **(J) CIRC. LIGHT EXP. APA** | F (5, 420) = 1.20 | | P = 0.30 | | F (1, 84) = 0.61 | P = 0.43 | | **F (5, 420) = 38.12** | **P < 0.0001** | |
| **(L) CIRC. LIGHT EXP. RA** | **F (5, 620) = 4.46** | | **P = 0.0005** | | **F (1, 124) = 5.7** | **P = 0.01** | | **F (5, 620) = 39.90** | **P < 0.0001** | |
| **(M) CIRC. TEMP. APA** | **F (5, 420) = 2.85** | | **P = 0.01** | | **F (1, 84) = 15.22** | **P = 0.0002** | | **F (5, 420) = 40.65** | **P < 0.0001** | |
| **(N) CIRC. TEMP. RA** | F (5, 620) = 1.54 | | P = 0.17 | | **F (1, 124) = 4.41** | **P = 0.03** | | **F (5, 620) = 55.77** | **P < 0.0001** | |
| **(M) CIRC. ACCELERATION APA** | **F (5, 420) = 8.25** | | **P < 0.0001** | | **F (1, 84) = 6.57** | **P = 0.01** | | **F (5, 420) = 83.40** | **P < 0.0001** | |
| **(O) CIRC. ACCELERATION RA** | F (5, 620) = 1.29 | | P = 0.26 | | F (1, 124) = 0.07 | P = 0.78 | | **F (5, 620) = 95.95** | **P < 0.0001** | |
| **Figure 1** | **INTERACTION** | | | | **GROUP FACTOR** | | | **ROUTIN/HABITS FACTOR** | | |
|  | F | | p | | F | p | | F | p | |
| **(A) ACROPHASE T. MOVEMENT** | F (1, 208) = 0.66 | | P = 0.41 | | **F (1, 208) = 6.41** | **P = 0.01** | | F (1, 208) = 3.29 | P = 0.07 | |
| **(B) ACROPHASE TEMPERATURE** | F (1, 208) = 1.99 | | P = 0.15 | | F (1, 208) = 1.92 | P = 0.16 | | F (1, 208) = 0.08 | P = 0.76 | |
| **(C) ACROPHASE LIGHT EXPOSURE** | F (1, 208) = 3.24 | | P = 0.07 | | F (1, 208) = 0.27 | P = 0.6 | | F (1, 208) = 0.18 | P = 0.66 | |
| **(D) ACROPHASE ACCELERATION** | F (1, 208) = 1.13 | | P = 0.28 | | **F (1, 208) = 4.78** | **P = 0.02** | | F (1, 208) = 1.76 | P = 0.18 | |
| **Table 2** | **INTERACTION** | | | **GROUP FACTOR** | | | | **ROUTIN/HABITS FACTOR** | | |
|  | F | p | | F | | | p | F | | p |
| **(A) TOTAL ACTIVITY (24H-T.MOV)** | F (1, 208) = 0.60 | P = 0.43 | | F (1, 208) = 0.009 | | | P = 0.92 | **F (1, 208) = 6.87** | | **P = 0.009** |
| **(B) TOTAL ACTIVITY (24H-A-G)** | F (1, 208) = 1.66 | P = 0.19 | | **F (1, 208) = 11.00** | | | **P = 0.001** | F (1, 208) = 2.94 | | P = 0.08 |
| **(C) AUC (A-G)** | F (1, 208) = 2.42 | P = 0.12 | | **F (1, 208) = 11.05** | | | **P = 0.001** | F (1, 208) = 3.11 | | P = 0.07 |
| **(D) ACTIVITY PERIOD (A-G)** | F (1, 208) = 2.07 | P = 0.15 | | **F (1, 208) = 9.44** | | | **P = 0.002** | F (1, 208) = 0.01 | | P = 0.90 |
| **(E) REST PERIOD (A-G)** | F (1, 208) = 1.41 | P = 0.23 | | **F (1, 208) = 6.87** | | | **P = 0.009** | F (1, 208) = 1.16 | | P = 0.28 |
| **(F) HIGH ACTIVITY (MIN A-G) >80** | F (1, 208) = 0.15 | P = 0.69 | | **F (1, 208) = 19.50** | | | **P < 0.0001** | F (1, 208) = 0.08 | | P = 0.77 |
| **(G) LOW ACTIVITY (MIN A-G) <5** | F (1, 208) = 0.30 | P = 0.58 | | **F (1, 208) = 5.17** | | | **P = 0.02** | F (1, 208) = 0.007 | | P = 0.92 |
| **(H) BED TIME** | F (1, 208) = 1.44 | P = 0.23 | | **F (1, 208) = 9.47** | | | **P = 0.002** | **F (1, 208) = 10.06** | | **P = 0.001** |
| **(I) WAKE UP TIME** | F (1, 208) = 2.41 | P = 0.12 | | **F (1, 208) = 13.10** | | | **P = 0.0004** | **F (1, 208) = 48.17** | | **P < 0.0001** |
| **(J) RESTING (MIN)** | F (1, 208) = 0.44 | P = 0.50 | | F (1, 208) = 2.37 | | | P = 0.12 | **F (1, 208) = 22.66** | | **P < 0.0001** |
| **(K) LUXES IN ACTIVITY PERIOD** | F (1, 208) = 1.61 | P = 0.20 | | **F (1, 208) = 4.77** | | | **P = 0.03** | **F (1, 208) = 8.35** | | **P = 0.004** |
| **(L) LIGHT AT NIGHT (MIN) >5 (12H)** | F (1, 208) = 0.21 | P = 0.64 | | **F (1, 208) = 10.11** | | | **P = 0.001** | **F (1, 208) = 9.41** | | **P = 0.002** |
| **(M) TEMPERATURE 24H** | F (1, 208) = 0.17 | P = 0.67 | | **F (1, 208) = 9.37** | | | **P = 0.002** | F (1, 208) = 0.78 | | P = 0.37 |
| **(N) TEMPERATURE AT DAY** | F (1, 208) = 1.67 | P = 0.19 | | **F (1, 208) = 9.25** | | | **P = 0.002** | **F (1, 208) = 13.21** | | **P = 0.0004** |
| **(O) TEMPERATURE AT NIGHT** | F (1, 208) = 0.06 | P = 0.79 | | F (1, 208) = 1.11 | | | P = 0.29 | F (1, 208) = 0.94 | | P = 0.33 |
