## supplementary table 5 for "Healthy behaviors and circadian patterns determined by actigraphy in researchers and administrative personnel as protective factors against metabolic disease and obesity"

**Supplementary Table 5**. Statistical data from Figure 1. Bold numbers indicates significant differences.

| **Supplementary**  **Table 5** | **ADMINISTRATIVE PERSONNEL** | | **RESEARCHERS** | | t | df | p |
| --- | --- | --- | --- | --- | --- | --- | --- |
|  | mean | sem | mean | sem |  |  |  |
| **SJL (MIN)** | **80.3** | **6.95** | 44.84 | 5.2 | **4.15** | **148** | **<0.0001** |
| **T. MOVEMENT** | 33.62 | 16.67 | 17.83 | 11.5 | 0.8 | 148 | 0.42 |
| **TEMPERATURE** | 12.23 | 27.67 | 86.53 | 33.87 | 1.57 | 148 | 0.11 |
| **LIGHT EXPOSURE** | **-60.72** | **21.49** | 26.31 | 15.94 | **3.31** | **148** | **0.001** |
| **ACCELERATION** | 31.28 | 17.57 | 5.42 | 13.66 | 1.17 | 148 | 0.24 |
